# European Health Regulations Reduce Registry-Based Research

**DOI:** 10.1101/2024.03.20.24304569

**Authors:** Oscar Brück, Enni Sanmark, Ville Ponkilainen, Alexander Bützow, Aleksi Reito, Joonas H. Kauppila, Ilari Kuitunen

## Abstract

The European Health Data Space regulation (EHDS) has been proposed to harmonize health data processing. Given its parallels with the Act on Secondary Use of Health and Social Data (Secondary Use Act) implemented in Finland in 2020, this study examines the consequences of heightened privacy constraints on registry-based medical research. Between 2020 and 2023, a median of 5.5% fewer data permits were approved annually by Finnish university hospitals. Based on linear regression modelling, we estimated a reduction of 46.9% in new data permits nationally in 2023 compared to the expected count. Similar changes were not observed in other medical research types highlighting the consequences of excessive data privacy laws on registry-based medical research.

## INTRODUCTION

Registry-based medical research forms an integral pillar of modern healthcare. Medical registries are invaluable resources to generate evidence-based medical guidelines and develop innovations such as effective diagnostic and monitoring tools. However, this progress has been subjected to growing regulatory burden in the European Union (EU), particularly in the realm of data privacy and security. The General Data Protection Regulation (GDPR), implemented in 2018, has strengthened personal data protection rights by providing individuals with greater control over their data, and imposing stricter obligations on data controllers and processors. Currently, the European Health Data Space regulation (EHDS) is under preparation to establish a common framework for the governance and sharing of health data across the European Union. While this initiative holds promise for facilitating cross-border access to health data, it also introduces additional layers of bureaucracy and potential delays in research and innovation raising questions of its effectiveness^1^.

To complement the GDPR, the Ministry of Social Affairs and Health introduced the Act on the Secondary Use of Health and Social Data (Secondary Use Act) in Finland in 2019 and implemented it into use in April 2020. Its primary purpose was to facilitate the effective and safe processing of personal social and health data for secondary uses, such as research, policymaking, and healthcare management, in contrast to primary uses, such as provision of healthcare services. Individual public health care entities, such as university hospitals, retained the right to grant data permits to their own data in single-registry studies. Instead, Findata was established as the centralized data permit authority, *i.e*. a one-stop-agency granting data permits for secondary use, combining data from private healthcare providers, the national Patient Data Repository, or a combination of multiple social and health registries, with multiple other tasks, such as defining policies for safe data processing.

Despite these positive objectives, two independent surveys conducted in 2021 highlighted the discontent of the research community. The first study was directed to medical doctors and revealed that out of 430 responders 79% experienced the data permit process to have become more complex than before, 55% that research projects were delayed, and 64% that research costs had increased^2^. Due to these issues, 42% of responders reported that they had not initiated a study^2^. Similar results were reported in a second survey commissioned by the Ministry of Social Affairs and Health where responders (n=260) evaluated that the Secondary Use Act had neither simplified the application process nor the combination of multiple registries^3^.

While the EHDS regulation is expected to harmonize the principles of health data processing in the EU, there is little retrospective evidence on the impact of increasing privacy regulations on registry-based social and medical research. Given its considerable overlap with the Secondary Use Act, we sought to evaluate the impact of the regulation on registry-based research by examining data permit counts before and after the implementation of the Act.

## METHODS

### Data permits

There are five university hospitals in Finland, located in Helsinki, Tampere, Kuopio, Turku, and Oulu. When referencing these cities, we are specifically alluding to the respective university hospitals. University hospitals serve as regional hubs for specialized care and providing comprehensive medical services to their respective catchment areas. The hospitals are well-equipped for registry-based research due to their advanced IT infrastructure and larger patient populations, enabling them to effectively collect, manage, and analyze large-scale health data both for primary and secondary purposes.

After entry into force of the Secondary Use Act, both a study permit and data permit were required to conduct registry-based research projects in Finland. In this study, we employ the term ‘data permit’ as university hospitals can grant both study and data permits but Findata can grant only data permits.

In January 2024, we solicited counts of new data permits for registry-based research, specifically involving university hospital registries, from the research departments of all five university hospitals (Helsinki, Turku, Tampere, Oulu, and Kuopio). To account for variations in research funding and regulatory changes, we also requested data on study permits for medical research involving subjects, human tissue, or medical devices. Due to varying archiving practices, data from 2016-2023 were available from Tampere, Oulu, Kuopio, and Helsinki. As data was accessible only from 2020-2023, Turku was excluded in analyses requiring data prior to 2020.

We also solicited counts of new data permit (n=375) involving clinical data from university hospitals and granted by Findata in 2020-2023. Recognizing that Findata-approved permits cover data from 1-5 hospitals, we integrated details on the annual mean count of university hospitals covered by these permits (mean 2.2-2.6 in 2021-2023).

### Statistical analysis

We fitted univariate linear regression analyses using data permit counts as covariate and year as predictor and examined the slopes of the curves by extracting the coefficient. We used the Mann-Kendall test to estimate trend over time. Based on prior findings, we reasoned to test only whether the registry-based study count had decreased since its implementation with the one-sided Mann-Kendall test^2,3^. Elsewhere, we applied the two-sided tests. We performed statistical analyses and visualizations with R 4.0 using the packages base, sf, mapsFinland, ggplot2.

## RESULTS

In 2020-2023, 1768 registry-based research data permits have been granted by university hospitals (Fig. 1a). Most of these (n=595) were approved by Helsinki, followed by Tampere (n=367), Turku (n=355), Oulu (n=231), and Kuopio (n=220). Following a stable period between 2016 and 2019 (median 517, range 497-573; Fig. 1b), new data permit counts decreased rapidly across hospitals (tau -1, p=0.042, one-sided Mann-Kendall test). Compared to 2019, there was a median decrease of 22.4 (range 9.7-31.7) data permits per year, representing an annual median reduction of 5.5% (range 3.4-10.5%). During the same time, 375 registry-based data permits were approved by Findata (Fig. 1b). For unclear reasons, the rate of Findata-approved data permits also decreased from 141 to 99 (29.8%) in 2023 (Fig. 1b).

**Figure 1.**
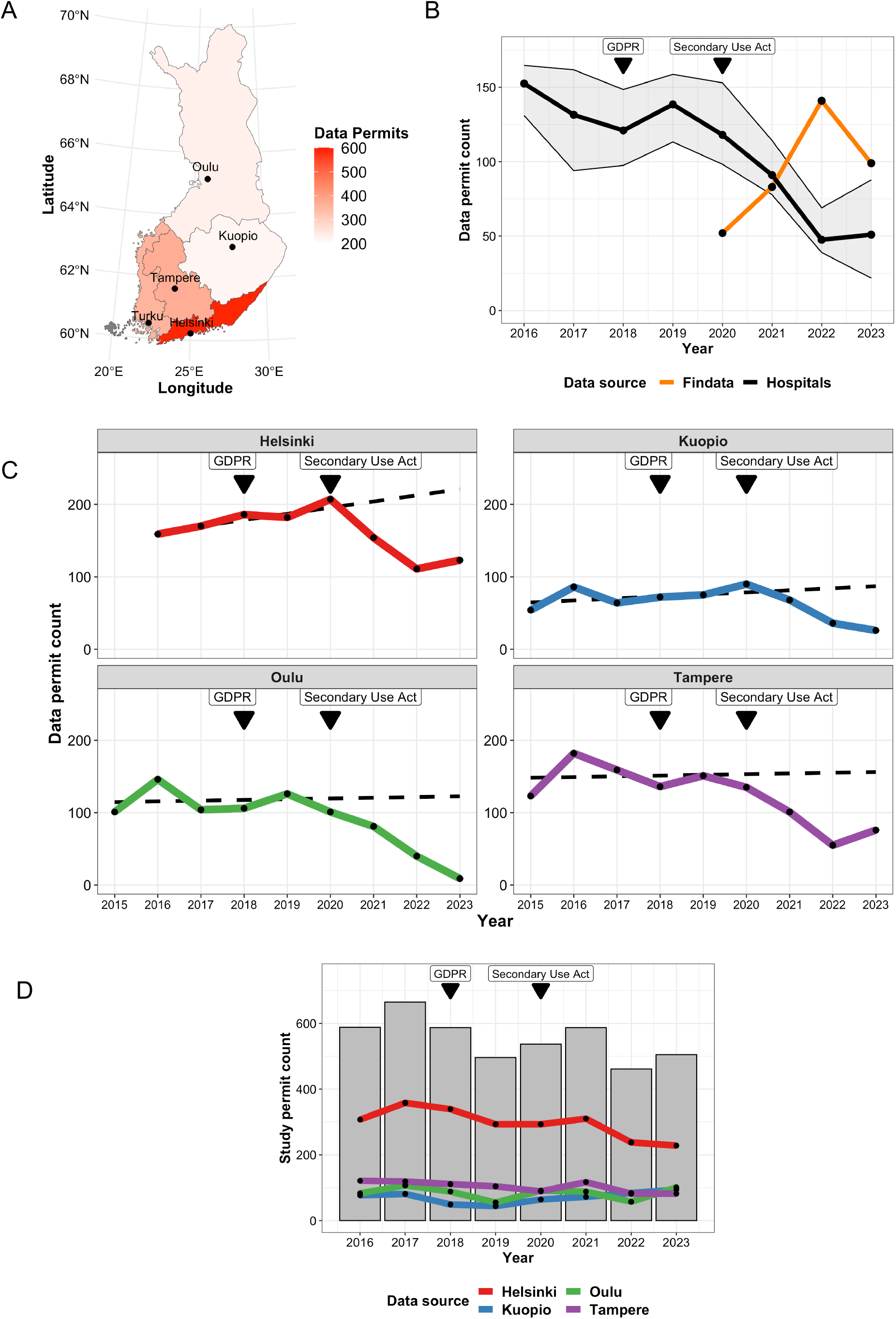
Registry-based research in Finland. (a) Map of Finland demonstrating cities (points) with a university hospital and their associated healthcare regions. (b) Line plots depicting the median counts of registry data permits approved by four Finnish university hospitals (black line) and data collections transferred by Findata (orange line). The gray-shaded area represents the 25-75% interquartile range of permit counts from university hospitals. (c) Line plots of median counts of approved registry data permits by distinct Finnish university hospitals in 2015-2023. These are accompanied by fitted regression curves predicting permit counts based on trends before the Secondary Use Act in 2015-2019 (dashed lines). (d) Data permit counts for other types than registry-based research are illustrated both individually (line plots) and cumulatively (bar plots) for each university hospital.

Next, we examined data permit counts approved by university hospitals before and after (2016-2017 vs. 2018-2019) the implementation of the GDPR. An annual median of 152 new registry research permits were approved pre-GDPR (Fig. 1b). While the corresponding median in the following two years was 131, the count sharply increased from 121 in 2018 to 138 permits in 2019 implying the GDPR had no conclusive impact on data permit counts (Fig. 1b).

To estimate the decline in data permits, we examined the era before the implementation of the Secondary Use Act. From 2015 to 2019, a median of 1.9 (range 1.0-7.3) additional data permits were approved corresponding to a 0.70% (range 0.20-1.3%) yearly accumulation. Assuming that similar progress would have continued until 2023, we fitted a linear regression curve for each hospital and predicted that a total of 586.8 new data permits would have been granted in 2023 (Fig. 1c). The figure contrasts with the 234 new data permits actually approved. Given that a portion of multi-center permits have been directed to Findata and as Turku was excluded from the analyses due to missing data, we included data permits accorded by Findata in 2023 (n=99) multiplied by 78.3% reflecting the percentage of data permits approved by other university hospitals than Turku in 2023. The estimated reduction amounted to 275.3 (= 586.8 – 234.0 – (99.0 × 0.78)) data permits corresponding to a relative reduction of 314.2/586.8 = 46.9%.

To exclude other confounding factors, we examined the data permit counts for other types of medical research (Fig. 1d). While the proportion of approved permits decreased in 2016-2023 (tau -0.54, p=0.061, two-sided Mann-Kendall test), the changes did not coincide with neither the enactment of the GDPR nor of the Secondary Use Act.

## DISCUSSION

This retrospective observational cohort study demonstrates the devastating impact of the Secondary Use Act on registry-based studies only three years after its implementation. The findings sharply contrast with the general positive attitude towards secondary use of personal data in medical research^4^.

This study has several implications for efforts to understand the potential impact of the EHDS regulation on research and innovations. First, while registry-based research could be previously performed with a minimal budget, solid funding has become a necessity following the enactment of the Secondary Use Act. Primarily data collections, but also application fees for submitting a study plan, adding researchers to a valid permit, and processing data in secure cloud-based computing instances accumulate significant research costs.

Secondly, stringent data privacy laws may hinder individual countries from participating in multicenter registry studies, and potentially impede global health by slowing down responses to pandemics, such as COVID-19^5^. Finland has a long tradition of nationwide healthcare registries, such as the care registry for specialized healthcare, the cancer registry, and implant registries, which all have been acknowledged for their coverage and quality^6–8^. Integrating Finnish registry holders in international studies is possible only if data is processed in a computing environment that have been audited to meet the legal requirements. Currently, only nine such environments are eligible, and all are based in Finland, implying that international registry studies would require transferring all data to one of these environments^9^.

The main source of error in this study stems from the prediction of new data permits in 2023 in a scenario without the Secondary Use Act. However, we anticipate that registry-based research would have gained even more popularity than before, given substantial investments in healthcare information technology infrastructures facilitating the release of electronic health records to cloud-based computing instances. Thus, many researchers experienced that the regulatory environment hampered not only conventional registry-based studies but also the promising progress towards automatic computer-assisted data curation and disease phenotyping^2,10^.

In conclusion, the results emphasize the need to balance between effective and data secure research. Medical researchers should be involved in planning, interpreting and assessing health data regulations. Besides their complexity, the cumulative effect of European and national regulations has created a challenging environment for medical researchers. Instead of improving patient privacy rights, increased administrative work and excessive technology requirements to ensure security may delay research projects and accumulate steeping costs. Ultimately, the regulatory burden may turn against its objectives and impede progress in patient care^11^.

## ACKNOWLEDGMENTS

The author wishes to thank Susanna Laurén (Turku University Hospital), Jaana Jäppinen (Helsinki University Hospital), Minna Mäkiniemi (Oulu University Hospital), Satu Ranta (Tampere University Hospital), and Tuomas Selander (Kuopio University Hospital) for providing the data for the data permit counts for their university hospital. In addition, we would like to thank Antti Piirainen, Maari Parkkinen, and Johanna Seppänen (Findata) for providing information on the registry holders of Findata data permits as well as for insightful comments. This study was supported by research funding from the Helsinki University Hospital, Finnish Cancer Foundation, Research Council of Finland, and the University of Helsinki.

## DATA AVAILABILITY STATEMENT

Source data and codes are available at https://github.com/obruck/Nav-Reg-Res.

## AUTHORS’ CONTRIBUTIONS

Conception and design: O.B. Collection and assembly of data: O.B. Data analysis: O.B. Data interpretation: All authors. Manuscript writing: O.B. Manuscript editing: All authors. Final approval of manuscript: All authors.

## REFERENCES

1 Marelli L, Stevens M, Sharon T, et al. The European health data space: Too big to succeed? Health Policy 2023; 135: 104861.

2 Reito A, Sanmark E, Tuovinen T, et al. Enabler or suppressor? – Survey on the effects of the Act on the Secondary Use of Health and Social Data on medical research. Suom Lääkärilehti 2022; 77. https://www.laakarilehti.fi/e30589.

3 Koiste V, Siranko H. Preliminary investigation on the impact of the Act on the secondary use of health and social data in research, development and innovation activities and in teaching. Gesund Partners, 2022.

4 Richter G, Borzikowsky C, Lesch W, et al. Secondary research use of personal medical data: attitudes from patient and population surveys in The Netherlands and Germany. European Journal of Human Genetics 2021; 29: 495–502.

5 Bak M, Madai VI, Fritzsche M-C, Mayrhofer MTh, McLennan S. You Can’t Have AI Both Ways: Balancing Health Data Privacy and Access Fairly. Frontiers in Genetics 2022; 13. https://www.frontiersin.org/articles/10.3389/fgene.2022.929453.

6 Mäkelä KT, Furnes O, Hallan G, et al. The benefits of collaboration: the Nordic Arthroplasty Register Association. EFORT Open Reviews 2019; 4: 391–400.

7 Sund R. Quality of the Finnish Hospital Discharge Register: A systematic review. Scandinavian Journal of Public Health 2012; 40: 505–15.

8 Leinonen MK, Miettinen J, Heikkinen S, Pitkäniemi J, Malila N. Quality measures of the population-based Finnish Cancer Registry indicate sound data quality for solid malignant tumours. European Journal of Cancer 2017; 77: 31–9.

9 National Supervisory Authority for Welfare and Health. Database on secondary-use environments (TOINI). 2023.

10 Brück O, Lallukka-Brück S, Hohtari H, et al. Machine Learning of Bone Marrow Histopathology Identifies Genetic and Clinical Determinants in MDS Patients. Blood Cancer Discovery 2021; 2: 238–49.

11 Huusko J, Kinnunen U-M, Saranto K. Medical device regulation (MDR) in health technology enterprises – perspectives of managers and regulatory professionals. BMC Health Services Research 2023; 23: 310.

